# Pre-bed and nighttime screen use, beyond daily total, is inversely associated with sleep quality: a longitudinal study of 350,600 nights

**DOI:** 10.64898/2026.05.08.26352708

**Authors:** Kanika Gupta, Nihav Dhawale, Aditi Shanmugam, Vinayak Narasimhan

**Affiliations:** Ultrahuman Healthcare Pvt Ltd, Bangalore, India

**Keywords:** screen time, sleep quality, pre-bed screen exposure, nighttime screen use, wearable, smartphone use, blood biomarkers

## Abstract

Sleep is fundamental to metabolic regulation, cognitive performance, immune function, and cardiovascular health, and evening screen exposure is widely proposed as a behavioural contributor whose adult evidence base remains thin. Here we analyze 350,600 paired screen-day and following-night observations from 3,086 Ultrahuman Ring AIR adult users. Sleep quality was assessed via the ring’s composite sleep score, derived from heart-rate variability, nightly movement, and skin temperature. At the user level, users in the highest screen-time quintile had lower sleep scores (Cohen’s d = −0.30), shorter sleep duration (d = −0.25), and lower sleep efficiency (d = −0.14) than the lowest screen-time quintile (all q ≤ 0.005). Further, 45+ min of screen use in the last hour before bed was associated with mean sleep scores at the bottom of the cohort range, whereas the same dose 4–5 hours earlier showed no detectable cost, so the timing of screen use, not just its total, mattered. We next asked whether the way users distribute their screen time across the 24 hours, independent of total dose, separates users by sleep outcome. K-means clustering on 24-hour screen-use profiles identified three phenotypes: Daytime Peakers (DP), Late-Night Users (LNU), and Round-the-Clock Users (RCU), distinguished primarily by their nighttime share of 24-h screen use (DP 8.2%, LNU 16.9%, RCU 29.3%). Despite comparable total daily screen time, the phenotype gap in mean sleep score between DP (75.2 ± 0.3 SEM) and RCU (66.7 ± 0.6) was 8.5 points. We further identified users who transitioned phenotypes across four sequential quarters of follow-up; in this longitudinal subcohort, the magnitude of sleep-score change tracked the magnitude of the behavioural shift, with DP → LNU transitioners declining by 5.16 ± 0.94 points and LNU → RCU transitioners by 4.79 ± 1.87 points (both p < 0.05). Together, these findings position the temporal distribution of screen use, alongside its daily total and its concentration immediately before bed, as a behavioural correlate of objectively measured sleep quality in adults.

## Introduction

Sleep is fundamental to metabolic regulation, cognitive performance, immune function, and cardiovascular health [1–3]. Insufficient or poor-quality sleep has been associated with increased risk of obesity, type 2 diabetes, depression, and all-cause mortality [1,4], and an estimated one-third of adults in industrialized nations sleep less than the recommended seven hours [5,6]. Evening screen time has been proposed as a behavioral contributor to impaired sleep [7–9], yet the adult evidence base linking screen time to objectively measured sleep in real-world populations remains thin, for specific, methodological reasons we discuss next.

Most epidemiological studies of screen time and sleep rely on self-reported screen exposure or short-duration actigraphy-based sleep measures [14]. A 2025 systematic review and meta-analysis across 21 cohort studies found that few studies used device-based screen-time measurement, and those that did were generally limited in sample size or duration [15]. A 2024 National Sleep Foundation consensus statement independently highlighted the need for studies combining passively sensed screen exposure with objective sleep assessment as a major methodological priority [16]. Recent studies using objective smartphone measures have focused on adolescent populations [17,18], leaving a notable gap in adult cohorts.

Here we combine passively sensed screen time with wearable-measured sleep in 3,086 adults across 350,600 matched nights. We test whether screen use in the hours preceding sleep is associated with reduced sleep quality and whether distinct temporal patterns of 24-hour screen use map onto different sleep outcomes.

## Results

### Daily screen time is inversely associated with sleep quality

In the matched cohort of 3,086 Ring AIR users (350,600 matched screen-sleep observation days; cohort assembly and disposition in Methods and **Table S1**), we asked whether average daily screen exposure was associated with sleep quality.

Per-user mean sleep score declined linearly with mean daily screen time across the matched cohort: the cross-sectional regression had slope −0.67 sleep-score points per additional hour of daily screen time (Pearson r = −0.108, Spearman ρ = −0.123, both p < 0.001; **Figure 1A** solid line). The cohort mean daily screen time was 2.97 hours per day (median 2.85 h; histogram bars in **Figure 1A**). Stratifying users into daily-screen-time quintiles (616–618 users per quintile) and ordering across quintiles confirmed a monotone trend in mean sleep score (Jonckheere–Terpstra z = −6.98, p < 0.001).

**Figure 1.**
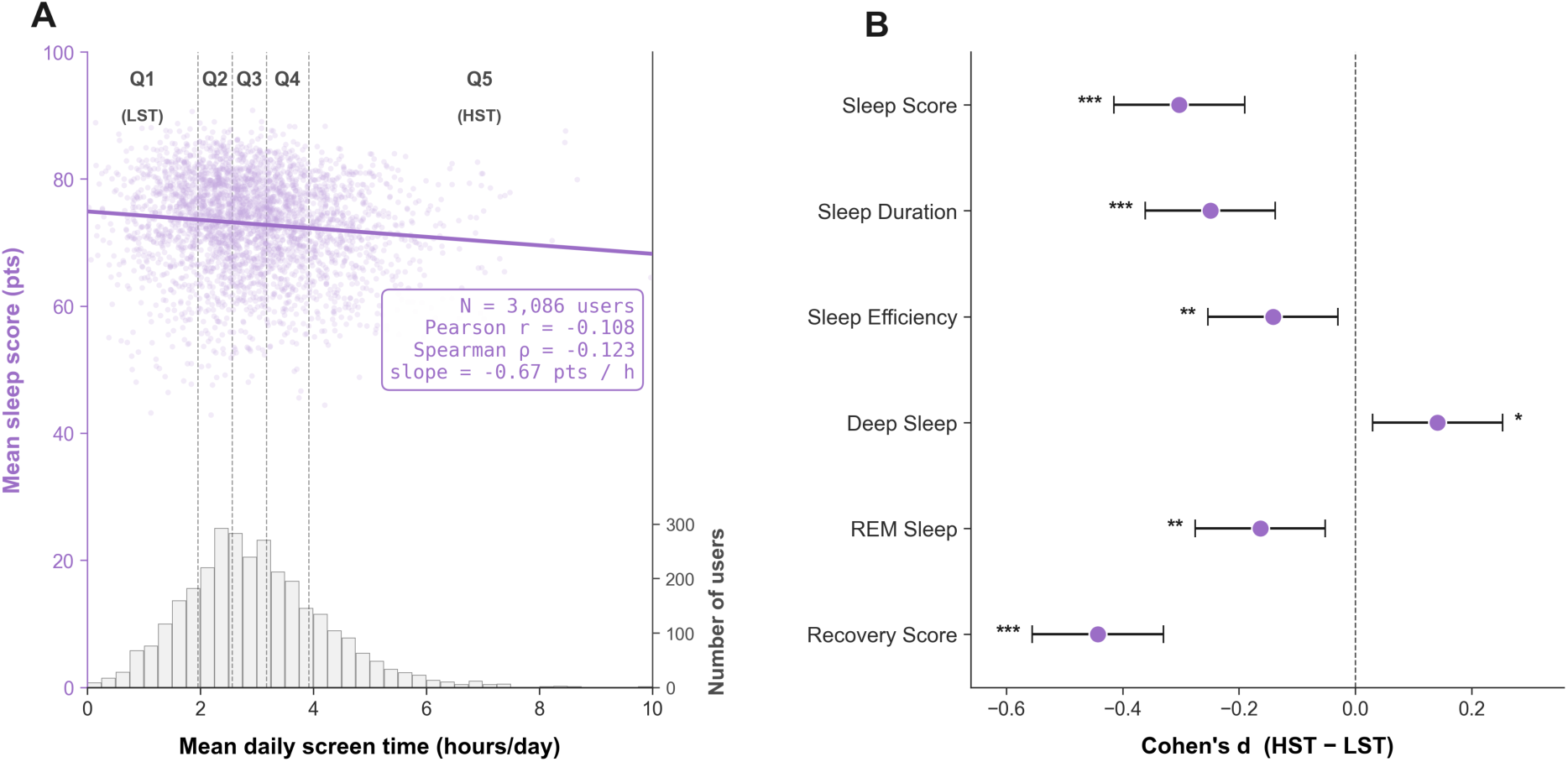
Daily screen time is inversely associated with sleep quality. **(A)** Per-user mean sleep score (left axis, points) versus mean daily screen time (x-axis, hours/day) for the matched cohort (N = 3,086; one purple dot per user). Solid black line = ordinary least-squares linear fit (slope = −0.67 sleep-score points per additional hour of daily screen time); shaded band = 95% confidence interval. Vertical dashed lines and Q1–Q5 labels at the top of the panel mark the four daily-screen-time quintile cut-points used to define the HST and LST groups in Panel B. Lavender bars at the bottom of the panel are a histogram of users-per-bin (right axis, ‘Number of users’). **(B)** Forest plot of HST (Q5; N = 617) vs LST (Q1; N = 618) effect sizes across six user-level sleep architecture metrics. Each point is Cohen’s d (HST − LST); horizontal lines are 95% confidence intervals (large-sample SE). Negative d = HST worse than LST. Asterisks reflect Benjamini–Hochberg FDR-6 correction across the six tests applied to the per-metric test-of-record (* q < 0.05, ** q < 0.01, *** q < 0.001). Per-metric values are reported in **Table 1** and the Results.

**Table 1.** Sleep architecture metrics and demographics for the highest (HST, Q5) vs lowest (LST, Q1) daily-screen-time quintile. Three sub-tables. **(A)** User-level summary (LST N = 618; HST N = 617): per-user mean of each metric, with LST mean, HST mean, Δ (HST − LST), test (Mann–Whitney U where the metric was non-normal, otherwise Welch’s t-test), effect size (rank-biserial r for MW-U; Cohen’s d for t-test), and p-value. **(B)** Night-level summary (LST n = 71,052; HST n = 68,029): same metrics and same test scaffold computed across all matched nights, used as a sensitivity check on the user-level summary. **(C)** Per-quintile demographics: number of users **(N)**, number of matched nights (n), age (mean ± SEM), BMI (mean ± SEM), sex split as count (% of quintile), and male-to-female ratio. Age and BMI compared with Mann–Whitney U; sex split compared with χ².

| Metric | LST Mean | HST Mean | Δ | Test | Effect | p |
| --- | --- | --- | --- | --- | --- | --- |
| Sleep Score | 73.5 | 71.2 | -2.3 | MW-U | r=-0.196 | p < 0.001 |
| Duration | 8.0 | 7.7 | -0.2 | MW-U | r=-0.150 | p < 0.001 |
| Deep % | 16.3 | 16.8 | +0.5 | MW-U | r=0.078 | p = 0.018 |
| REM % | 22.4 | 21.6 | -0.8 | t-test | d=0.164 | p = 0.004 |
| Efficiency | 88.4 | 87.9 | -0.6 | MW-U | r=-0.097 | p = 0.003 |
| Recovery | 69.4 | 66.7 | -2.7 | MW-U | r=-0.258 | p < 0.001 |

B Night-Level Summary (n = nights)
| Metric | LST Mean | HST Mean | Δ | Test | Effect | p |
| --- | --- | --- | --- | --- | --- | --- |
| Sleep Score | 73.7 | 71.4 | -2.3 | MW-U | r=-0.103 | p < 0.001 |
| Duration | 8.0 | 7.8 | -0.2 | MW-U | r=-0.071 | p < 0.001 |
| Deep % | 16.3 | 16.7 | +0.4 | MW-U | r=0.026 | p < 0.001 |
| REM % | 22.5 | 21.6 | -0.9 | MW-U | r=-0.053 | p < 0.001 |
| Efficiency | 88.5 | 87.9 | -0.6 | MW-U | r=-0.045 | p < 0.001 |
| Recovery | 69.5 | 66.8 | -2.7 | MW-U | r=-0.123 | p < 0.001 |

|  | Q1 (Low ST) | Q5 (High ST) | Test |
| --- | --- | --- | --- |
| N (users) | 618 | 615 |  |
| n (nights) | 71,052 | 68,029 |  |
| Age | 42.5 ± 0.5 | 37.8 ± 0.4 | MW-U: p < 0.001 |
| BMI | 27.2 ± 0.3 | 28.4 ± 0.3 | MW-U: p < 0.001 |
| Male | 454 (73%) | 407 (66%) | χ²: p < 0.001 |
| Female | 134 (22%) | 190 (31%) | χ²: p < 0.001 |
| M:F Ratio | 3.4:1 | 2.1:1 |  |

The deficit was not confined to the composite sleep score. Comparing the highest screen-time quintile (HST, Q5; N = 617) with the lowest (LST, Q1; N = 618) at the user level, HST users had worse outcomes than LST users on every component of sleep architecture we examined except deep sleep (**Figure 1B**; per-metric tests, effect sizes and FDR-corrected q-values in **Table 1**). Sleep duration was 0.25 h shorter in HST than LST (Cohen’s d = −0.25, q < 0.001), sleep efficiency was 0.6 percentage points lower (d = −0.14, q = 0.005), and REM share was 0.8 percentage points lower (d = −0.16, q = 0.005). Recovery Score, Ultrahuman Ring’s overnight autonomic-recovery index, distinct from the sleep-architecture metrics above, was 2.7 points lower in HST than LST (d = −0.44, q < 0.001), the largest effect we observed in this comparison. Deep sleep was higher in HST (d = +0.14, q = 0.018), the only metric showing a directional reversal. The lower sleep quality in HST is reflected across several sleep-architecture measures, not just one.

### The inverse screen-sleep association holds across age, sex, and weekday/weekend strata

The user-level analysis in **Figure 1** describes between-person variation. To test whether the same inverse association also holds within-person on a night-to-night basis and generalizes across demographic strata, we fit a linear mixed model with random user intercepts on 337,371 nights from 2,958 users with age and sex available (**Supplementary Figure S1**). The within-person fixed-effect slope was β = −0.482 sleep-score points per hour of daily screen time (95% CI −0.55 to −0.42, p < 0.001; **Figure S1D**), the same direction and similar magnitude as the between-person slope from **Figure 1A** (−0.67 pts/h). The intra-class correlation was ICC = 0.269 (27% of sleep-score variance between users), justifying the random-intercept structure. Reference categories for each moderator were age 30–39, male, and weekday; below, β denotes a stratum baseline shift relative to its reference (intercept term in points), and β_int denotes a slope shift (interaction with screen time in pts/h).

Age-stratified adjusted sleep-score curves (**Figure S1A**) confirmed that every age group carried a negative screen-time slope, with users under 30 starting from higher baseline sleep scores; the slope was slightly attenuated in users aged 50+ (β_int = +0.148 pts/h, p = 0.004; **Figure S1E**), without changing direction.

Sex showed the most consistent effect (**Figure S1B**): female users (N = 848) had higher adjusted sleep scores than males (N = 2,110) across the entire range of daily screen time (β = +2.41 pts; **Figure S1E**).

Weekday and weekend adjusted sleep-score curves were largely overlapping (**Figure S1C**). Weekend nights had a slightly lower baseline (β = −0.79 pts, p < 0.001; **Figure S1E**), but the day-type interaction with screen time was not significant (β_int = −0.027 pts/h, p = 0.328): the per-hour cost of screen time was not detectably different on weekends versus weekdays.

The full GLMM coefficient set is shown in **Figure S1D** (cohort-wide model fit: intercept, screen-time slope, ICC, and sample size) and **Figure S1E** (per-stratum baseline and slope shifts relative to each reference category).

### Pre-bed screen exposure is more strongly associated with sleep loss than total daily screen time

Daily total screen time established the user-level association, but the GLMM above showed that each additional hour of daily screen time carries only a small per-night effect (β = −0.482 pts/h). We therefore asked whether timing within the day, specifically the minutes immediately before sleep onset, carries a sharper signal than the daily total. Across the 3,086-user matched cohort, screen use in the night window (22:00–05:00) accounted for 15.4% of total daily screen time on average (median 14.2%; **Figure 2A**). A considerable portion of users concentrated their screen time in this window (832 of 3,086 allocated more than 20%; 188 of 3,086 more than 30%), so nighttime screen use is common in this cohort rather than a rare-tail behaviour.

**Figure 2.**
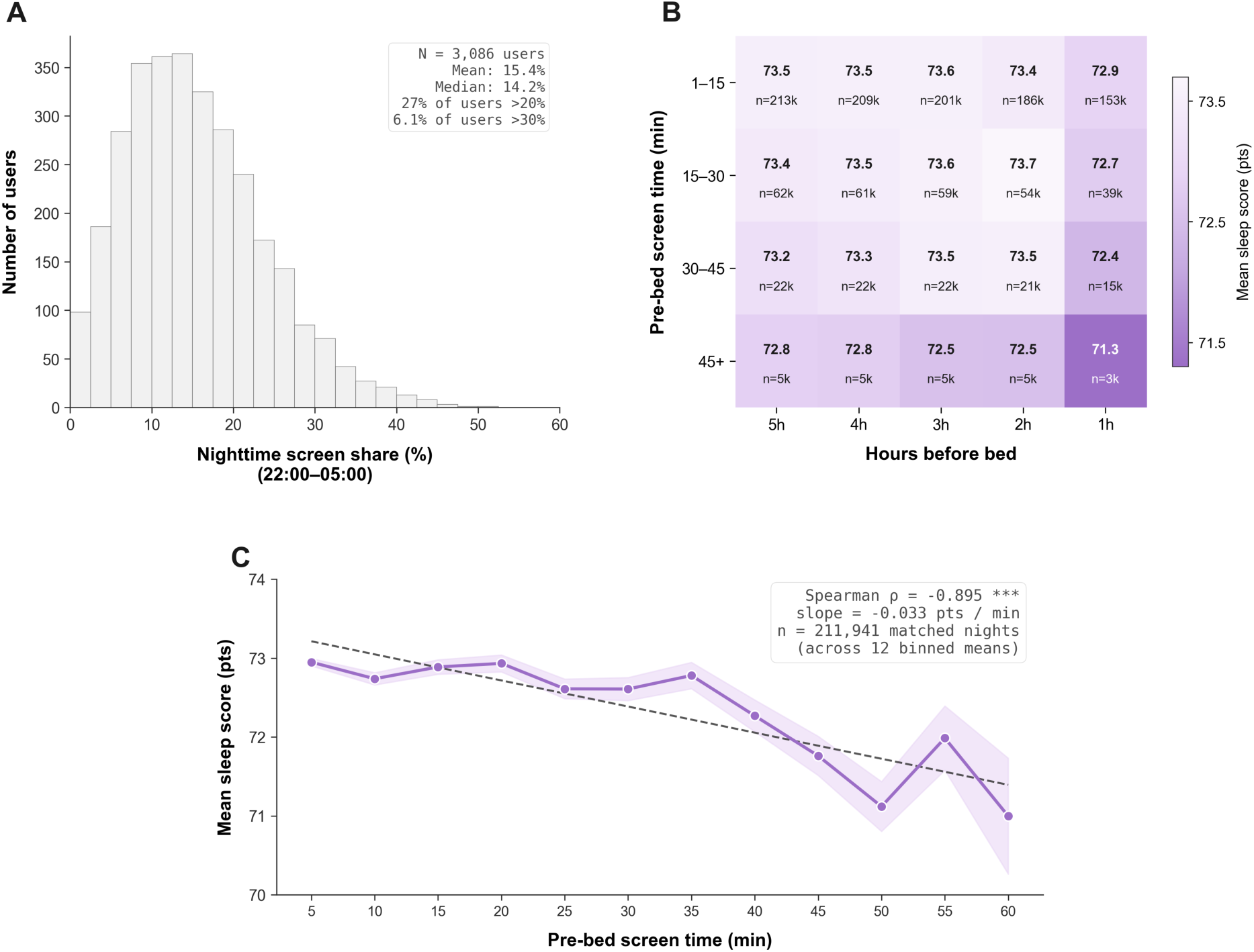
Pre-bed screen exposure is more strongly associated with sleep loss than total daily screen time. **(A)** Distribution of per-user nighttime screen share, the proportion of each user’s daily screen time falling between 22:00–05:00 (N = 3,086; mean = 15.4%, median = 14.2%; 27% of users allocate more than 20% of their screen time to the night window, 6.1% more than 30%). **(B)** Heatmap of mean sleep score per (hours before bed × pre-bed screen-time intensity) cell. Rows are non-overlapping pre-bed-minute buckets (1–15, 15–30, 30–45, 45+ minutes within the named clock-hour); columns are the five clock-hours immediately before each user’s habitual bedtime (1 h to 5 h before bed). Cell values are mean sleep score in points (grid mean = 73.08; cell range 71.34–73.71); n shown in each cell is the number of matched nights contributing to that cell (in thousands). The sequential white-to-purple colormap spans the displayed range; darker purple = lower mean sleep score, paler = higher. **(C)** Five-minute pre-bed dose-response: mean nightly sleep score (purple line, ± SEM band) as a function of total screen-time minutes in the hour immediately before bed, binned into twelve non-overlapping 5-minute bins (0–5, 5–10, …, 55–60). Dashed line is an OLS linear fit through the twelve bin means; the inset shows the Spearman rank correlation and linear slope computed across these binned means (ρ = −0.895, p < 0.001; slope = −0.033 pts/min).

Within the matched cohort, we mapped mean sleep score against two timing axes simultaneously: the clock-hour relative to each user’s habitual bedtime (1, 2, 3, 4, 5 hours before bed) and the amount of screen time accumulated within that hour (1–15, 15–30, 30–45, 45+ minutes). Each cell of the resulting heatmap (**Figure 2B**) is the mean sleep score across all matched nights whose screen-time minutes fall into that (hour-before-bed × intensity) bucket, on a sequential white-to-purple scale (cell range 71.3–73.7 sleep-score points). Two patterns are visible. First, within any single hour-before-bed column, mean sleep score decreased as pre-bed screen-time intensity increased: cells with 45+ minutes of screen time in the last clock-hour before bed sat at the bottom of the displayed range. Second, the same screen-time intensity was less penalising the further it was placed from bedtime: the column at 4–5 hours before bed showed almost no within-column gradient.

Narrowing to the single hour immediately before bed and resolving it into twelve 5-minute bins (**Figure 2C**; n = 211,941 matched nights with at least one minute of pre-bed screen time), mean sleep score declined steadily as pre-bed screen time increased. The dose-response across the twelve bin means was strongly inverse (Spearman ρ = −0.895, p < 0.001) and approximately linear (slope −0.033 sleep-score points per minute of pre-bed screen time). A user moving from no pre-bed screen use to a full hour of pre-bed screen use is therefore associated with an expected sleep-score decrement of roughly 2 points across the binned-mean trend; the same change in daily-total screen time, by contrast, is associated with under half a point at the user level (**Figure 1A**) and with the within-person GLMM slope above (β = −0.482 pts/h, ≈ −0.5 pts per added hour of daily screen time). Minute-for-minute, the pre-bed exposure tracks a larger sleep-score difference than the daily total.

### Phenotypes with similar daily screen time but more nighttime use sleep worse

The previous analyses considered total daily screen time and pre-bed timing in isolation. Users with similar daily totals can still distribute their screen time differently across the 24 hours, so we next asked whether stable patterns in that distribution separate users by sleep outcome independently of total dose. We applied K-means clustering to L1-normalized 24-hour screen-use profiles for the 3,086-user matched cohort (method and K-selection in Methods; cluster validation in **Supplementary Figure S2**); K = 3 was the working choice because it isolated a small but distinct phenotype maintaining elevated screen use through the night that K = 2 averaged into the broader population.

The three phenotypes (**Figure 3A**) were Daytime Peakers (DP; N = 1,161, 37.6% of the cohort), with screen use concentrated around 16:00 and the lowest screen use at night (nighttime share 8.2%); Late-Night Users (LNU; N = 1,477, 47.9%), peaking around 20:00 with moderate evening engagement (nighttime share 16.9%); and Round-the-Clock Users (RCU; N = 448, 14.5%), who maintained elevated screen use across all hours of the day (nighttime share 29.3% - more than triple DP and nearly double LNU).

**Figure 3.**
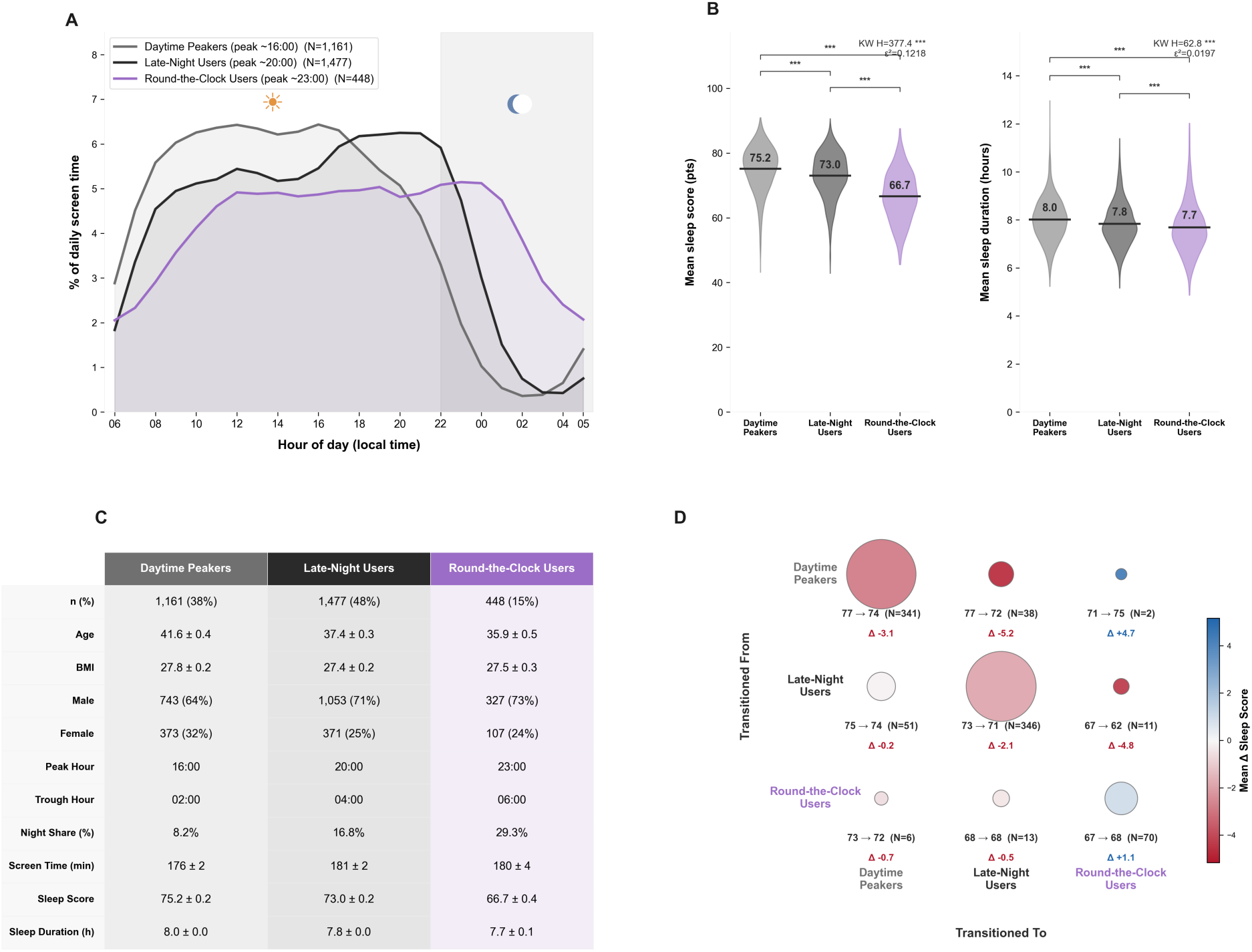
Higher nighttime share separates phenotypes with similar daily totals but different sleep outcomes. **(A)** Mean 24-hour screen-use profiles for the three K-means clusters (k = 3, L1-normalized), plotted in local time with 8 a.m. start: Daytime Peakers (n = 1,161, 38%, mean sleep score = 75.2); Late-Night Users (n = 1,477, 48%, mean sleep score = 73.0); Round-the-Clock Users (n = 448, 15%, mean sleep score = 66.7). **(B)** Sleep score and sleep duration distributions by cluster (Kruskal-Wallis followed by pairwise Mann-Whitney with Bonferroni correction; significance markers on the brackets are * p < 0.05, ** p < 0.01, *** p < 0.001). **(C)** Demographics summary table (n, age, BMI, gender split, peak/trough hour, mean daily screen time, sleep score, sleep duration) per cluster. **(D)** 3×3 transition matrix among phenotypes from baseline to follow-up (longitudinal cohort, n = 878 users with sustained cluster: 757 stayers, 121 changers); bubble area is proportional to n, color encodes mean Δ sleep score (red = decline, blue = improvement).

The three phenotypes differed in both sleep score and sleep duration (**Figure 3B**; per-cluster distributions in **Supplementary Figure S2**). DP had the highest mean sleep score (75.2 ± 0.3 SEM), followed by LNU (73.0 ± 0.3) and RCU (66.7 ± 0.6); the cluster-level Kruskal-Wallis was H = 377.4, p < 0.001, ε² = 0.12, and all three pairwise comparisons were significant (Mann-Whitney U with Bonferroni correction, all p < 0.001; rank-biserial r = 0.18 to 0.60). Total nightly sleep duration showed the same DP > LNU > RCU rank order (DP 8.0 ± 0.0 h, LNU 7.8 ± 0.0 h, RCU 7.7 ± 0.1 h; Kruskal-Wallis p < 0.001). Total daily screen time was comparable across phenotypes, so the sleep-score gap of 8.5 points between DP and RCU tracks the within-day distribution, in particular the nighttime share (8.2 → 16.9 → 29.3% across the three phenotypes). Because hard-assignment K-means places boundary users with intermediate profiles into RCU (**Supplementary Figure S2A**), the reported RCU mean sleep score (66.7) is a conservative estimate: restricting RCU to high-silhouette members would widen rather than narrow the DP–RCU gap. Demographic comparisons across the three phenotypes are shown in **Figure 3C** and tabulated in **Table S2**.

The cluster assignments above pool a user’s entire observation window into one 24-hour profile, so they cannot tell us whether a user is the same phenotype throughout, and whether the cross-sectional phenotype-sleep gap also tracks within-user behaviour change. To address this, we re-clustered each user-quarter independently (per-quarter clustering with centroid alignment to the cohort-level reference centroids; see Methods) on the 1,098 users with > 30 observation days in each of four sequential quarters spanning ≥ 12 months, and compared each user’s first-quarter cluster to the cluster they held across their last two recording quarters; 878 users had a stable last-two-quarter cluster and entered the transition analysis (**Figure 3D**). The interesting cell of the matrix is the off-diagonal: 121 users (13.8% of the 878-user transition cohort) sustained a phenotype change between baseline and follow-up, and the magnitude of the change in their mean sleep score scaled with the magnitude of the behavioural shift. Users moving DP → LNU (38 users) declined by 5.16 ± 0.94 sleep-score points (p < 0.001), users moving LNU → RCU (11 users) declined by 4.79 ± 1.87 points (p = 0.024), and the two users who made the largest behavioural shift in the taxonomy (DP → RCU) had a mean change of +4.72 points (Wilcoxon undefined at this small sample); transitions out of RCU (6 to 13 users per pair, all p > 0.5) did not reach significance, suggesting that the directional sleep-score effect of moving toward higher nighttime share is sharper than moving back toward lower. The remaining 757 users (86.2% of the transition cohort) were stayers; stayer-level sleep-score changes are reported alongside the off-diagonal transitions in **Table 2** and do not change the across-cohort phenotype gap reported above. Full transition statistics are in **Table 2**.

**Table 2.** Per-pair Wilcoxon signed-rank statistics for the 9-cell phenotype transition matrix supporting **Figure 3D**. For each (origin → destination) phenotype pair we report N, mean Δ sleep score (post − pre observation period), SEM, Wilcoxon W statistic, and p-value. Cells with N < 3 are reported as NA. Pairs supporting **Figure 3D** in the main text appear in the main-text body; this table provides the full matrix.

| From | To | N | Mean $\Delta \pm$ SEM | W statistic | p | Type |
| --- | --- | --- | --- | --- | --- | --- |
| Round-the-Clock Users | Round-the-Clock Users | 70 | +1.1 ± 0.9 | 1132 | p = 0.518 | Stayer |
| Round-the-Clock Users | Daytime Peakers | 6 | -0.7 ± 1.4 | 8 | p = 0.688 | Transitioner |
| Round-the-Clock Users | Late-Night Users | 13 | -0.5 ± 1.6 | 37 | p = 0.588 | Transitioner |
| Daytime Peakers | Round-the-Clock Users | 2 | +4.7 ± 7.0 | — | — | Transitioner |
| Daytime Peakers | Daytime Peakers | 341 | -3.1 ± 0.4 | 12788 | p < 0.001 | Stayer |
| Daytime Peakers | Late-Night Users | 38 | -5.2 ± 0.9 | 64 | p < 0.001 | Transitioner |
| Late-Night Users | Round-the-Clock Users | 11 | -4.8 ± 1.9 | 8 | p = 0.024 | Transitioner |
| Late-Night Users | Daytime Peakers | 51 | -0.2 ± 0.9 | 662 | p = 0.993 | Transitioner |
| Late-Night Users | Late-Night Users | 346 | -2.1 ± 0.3 | 18384 | p < 0.001 | Stayer |

Stayer rows (highlighted) represent users who maintained the same phenotype across observation period. Mean $\Delta$ = mean sleep-score change (last two quarters minus first quarter). SEM = standard error of the mean. Total: N = 878 users (757 stayers [86.2%], 121 transitioners [13.8%]).

### Blood biomarker profiles show directional trends across screen-use phenotypes

We next examined whether the behavioural phenotypes also carry detectable physiological signatures. We identified 65 users who had both a phenotype assignment and at least one measured analyte on a contemporaneous Ultrahuman Blood Vision panel (DP n = 14, LNU n = 36, RCU n = 15; demographic breakdown in **Table S2**; cohort-derivation detail in Methods). Across these 65 users, panel coverage varied (median 7 analytes per user), so the per-cutoff analytical N varies between 11 and 62; the per-cutoff and per-cluster cell counts are displayed alongside each row of **Supplementary Figure S3** so every odds ratio can be read against its own denominator. For each of 28 established clinical cutoffs across glycemic, lipid, hepatic, inflammatory, hormonal, and micronutrient domains we computed the proportion of users in each phenotype at or above the cutoff, plus an RCU-vs-DP odds ratio with Woolf 95% CI (**Supplementary Figure S3**).

No individual biomarker comparison reached statistical significance after FDR correction across the 28 tests, so the following are reported as directional trends, and we report each comparison as a count of users at or above the cutoff so the small denominators are visible in-text. In the glycemic domain, abnormality prevalence trended higher in DP than RCU for HbA1c ≥ 5.7% (9 of 14 DP, 15 of 35 LNU, 5 of 13 RCU; OR 0.35, 95% CI 0.07–1.66) and fasting glucose ≥ 100 mg/dL (5 of 12 DP, 8 of 34 LNU, 2 of 13 RCU; OR 0.26, 95% CI 0.04–1.69). Total cholesterol ≥ 200 mg/dL similarly trended higher in DP than RCU (8 of 14 DP, 14 of 35 LNU, 5 of 13 RCU; OR 0.47). The directional pattern flipped in the micronutrient and inflammatory domains: vitamin D deficiency (< 30 ng/mL) trended highest in RCU (6 of 8 RCU, 15 of 22 LNU, 3 of 8 DP; OR 5.0, 95% CI 0.58–42.80), and elevated hs-CRP (≥ 3.0 mg/L) similarly trended highest in RCU (4 of 10 RCU, 6 of 21 LNU, 2 of 9 DP; OR 2.33). The directionally consistent split, glycaemic/lipid burden concentrated in DP and inflammatory/micronutrient burden concentrated in RCU, sketches a phenotype-level physiological pattern that is consistent with the behavioural taxonomy: heaviest persistent screen engagement tracks low vitamin D and elevated systemic inflammation, directions that are mechanistically plausible given reduced outdoor light exposure and sleep-disruption-related inflammation.

A combined screen-time and biomarker cohort of 65 users is unusual at this scale, but the per-cluster, per-cutoff cell counts are small, so the confidence intervals on the RCU-vs-DP contrast are wide and no single cutoff carries inferential weight. Of note is the consistency of the directional pattern across two mechanistically distinct domains in users for whom we have simultaneous behavioural exposure, wearable sleep, and clinical biomarker data.

## Discussion

This study shows that pre-bed and nighttime screen use, beyond their daily total, are inversely associated with adult sleep quality at population scale. Three timing scales of screen-use exposure, the daily total, the minutes immediately before bed, and the 24-hour distribution, were examined in 3,086 adults contributing 350,600 paired screen-day and following-night observations.

Several mechanisms plausibly link evening screen exposure to impaired sleep, though our observational data cannot adjudicate between them. Light-emitting displays suppress melatonin secretion and delay circadian timing [10,11]. Engaging content sustains cognitive arousal, and late-night smartphone use has been shown to deplete next-day cognitive resources, mediated by sleep loss [12,13]. The relative contributions of light, content arousal, and displaced sleep opportunity to the dose-response we observe here cannot be separated without an experimental manipulation.

Pre-sleep screen exposure was a robust correlate of objectively measured sleep quality in this 3,086-user adult cohort. The pre-bed dose-response showed that sleep scores declined monotonically across the final hour before sleep (Spearman r = −0.895; −0.033 sleep-score points per minute), and the heatmap of screen use across the 5 hours preceding bed showed that the steepest decrements were concentrated at high screen-use durations closest to bedtime. Recent reviews and consensus statements have explicitly identified the lack of adult cohorts in which both screen exposure and sleep are measured objectively [15,16], and our findings address this gap by anchoring the screen–sleep relationship in objectively recorded behavior rather than recall.

The per-night dose-response (**Figure 2C**) and the user-level phenotype gap (**Figure 3B**) describe the same association at two timescales. On any single night, the marginal effect of an additional minute of pre-bed screen use on sleep score is small (−0.033 points/min, ∼2 points across the full last hour). What turns this small per-night slope into a substantial user-level gap is habituation: a user whose 24-hour profile concentrates substantial screen time in the late-night window night after night accumulates that small per-night effect across hundreds of nights, producing the 8.5-point sleep-score difference between Daytime Peakers (75.2) and Round-the-Clock Users (66.7) seen in **Figure 3B**. The bidirectional gradient in **Figure 1B** is consistent with a reinforcing cycle, and the observational design precludes causal inference, reverse causation, where individuals with poorer sleep engage in more evening screen use, remains plausible.

The screen-sleep association extended beyond the composite sleep score to multiple sleep architecture metrics. Users in the highest screen-time quintile showed shorter sleep duration, lower REM fraction, and notably lower next-day recovery scores. Recovery score showed the strongest user-level effect size (r = −0.258), suggesting that screen-time-related sleep disruption is most apparent in next-day physiological recovery, a dimension rarely measured in screen-time studies, in part because most prior cohorts lack the wearable infrastructure to compute it [19]. Our screen-time measurement was nonetheless limited to aggregate device interaction events; we could not distinguish content type, brightness, or active versus passive use, all of which may modulate sleep through blue-light vs cognitive-arousal pathways [10,11].

The three screen-use phenotypes, Daytime Peakers, Late-Night Users, and Round-the-Clock Users, revealed that the temporal distribution of screen use captures meaningful variation that aggregate daily totals miss. The three groups had only small differences in total daily screen time and sleep duration, yet their nighttime screen share differed more than threefold (8.2% in Daytime Peakers, 16.9% in Late-Night Users, and 29.3% in Round-the-Clock Users) and their sleep scores differed by up to 8.5 points (**Table S2**). A notable finding from the longitudinal subcohort was the 86% stability of phenotype assignment across four quarters: only 121 of 878 users with a stable cluster in their final two quarters changed phenotype across the observation window. This suggests these are durable behavioural patterns over the observed timescale; their responsiveness to active intervention remains a question for prospective work. Among the 121 transitioners, transitions toward higher-evening-use phenotypes were associated with substantial sleep-score declines (DP→LNU: −5.2, p < 0.001; LNU→RCU: −4.8, p = 0.02). The cohort recruited itself by purchasing the Ultrahuman Ring AIR and could be enriched for technology-engaged users; this convenience sampling structure should be considered when generalising our findings to the wider population.

The GLMM confirmed a negative slope of −0.482 sleep-score points per hour of daily screen time across the full cohort (p < 0.001). Female users had higher baseline sleep scores than males and a slightly attenuated screen-time penalty (β_int = +0.185 pts/h, p < 0.001), consistent with reported sex differences in sleep quality [22]. Users aged 50+ showed a slightly weaker association than younger groups, and our cohort adds to the limited literature on passively recorded screen time in adults aged 50+, where wearable sleep data have been particularly sparse, the size and direction of the age modification will need to be confirmed independently.

Past studies have shown that chronic short or disrupted sleep impairs insulin sensitivity [24], dysregulates the hypothalamic–pituitary–adrenal axis with consequences for cortisol rhythmicity and prolactin secretion [23], and elevates inflammatory markers including C-reactive protein [4]. Our biomarker trends are consistent with these mechanistic links and extend them toward a screen-time-anchored phenotype. Although the per-cell sample sizes are small (most contingency cells contain fewer than 12 users), the higher prevalence of vitamin D deficiency in Round-the-Clock Users (75% vs 38% in Daytime Peakers) is consistent with reduced outdoor light exposure, the principal source of cutaneous vitamin D synthesis [26], and the higher hs-CRP prevalence in Round-the-Clock Users (40% vs 22%) aligns with the sleep–inflammation link [4]. The inverse glycemic pattern (higher abnormality burden in Daytime Peakers) is harder to interpret without access to medical history, family history, supplementation status, or other biomarker covariates that would normally control for non-screen-time drivers of these clinical cutoffs.

Taken together, our findings position the temporal distribution of screen use, not just its daily total, as the behavioral correlate most strongly correlated with objectively measured sleep quality in adults. Combining passively sensed screen exposure with wearable-measured sleep architecture on a single platform let us trace this relationship from a behavioral correlate through sleep architecture.

## Methods

### Study design and participants

This retrospective observational cohort study analyzed data from users of the Ultrahuman Ring AIR who had valid screen-time recordings between 25 July 2024 and 5 March 2026. The full tabular flowchart for cohort derivation is in **Supplementary Table S1**; briefly: (1) the device-data starting set comprised all Ultrahuman Ring AIR users with valid screen-time recordings in this window; (2) we restricted to 3,333 users with ≥150 days of valid screen-time recording (longitudinal coverage filter); (3) of these, 3,086 users had ≥30 matched screen-sleep night pairs (each valid screen day paired with the following night’s sleep record) and form the primary analytical cohort (350,600 matched nights; supports **Figures 1, 2,** and S3); (4) the GLMM subcohort was further restricted to 2,958 users with non-missing age and sex (337,371 matched nights; supports **Figure S1**); (5) the HST/LST architecture-comparison subcohort comprised the 1,235 users in the lowest and highest daily-screen-time quintiles of cohort 02 (supports **Figure 1B** and **Table 1**); (6) the longitudinal phenotype-transition subcohort comprised 1,098 users with ≥30 days of valid data in each of four sequential quarters spanning ≥12 months (supports **Figure 3D** and **Table 2**); and (7) the biomarker subcohort comprised 91 users with both a contemporaneous Blood Vision panel and a phenotype assignment from cohort 02; of these, 65 had at least one measured analyte against the 28 clinical cutoffs examined and form the analytical biomarker subcohort (supports **Supplementary Figure S3** and **Table S2**). Each subcohort is fully nested within cohort 02 and is identified explicitly in the figure that uses it.

### Ethics statement

This was a real-world, retrospective, observational study based on data derived from Ultrahuman platform users, conducted in accordance with Ultrahuman’s <u>Terms of Use</u> and <u>Privacy Policy</u>, which permit the analysis of de-identified, aggregated data for scientific research. Participants consented through the onboarding process on the Ultrahuman platform and continued product use. No dietary, sleep, exercise, or other interventions were administered as part of this study: wearable-derived physiological signals and screen-time records were captured passively during routine product use, and blood biomarker results were obtained from contemporaneous Ultrahuman Blood Vision panels. All data were de-identified prior to analysis, and a separate team of analysts extracted the data, executed the computational pipelines, and reviewed the results to maintain analyst blinding.

### Screen-time measurement

Screen time was captured passively from participants’ paired Android smartphones through the Ultrahuman mobile application. Participants consented to share screen-time usage data with the application; the app recorded device interaction events including screen-on events, application foreground time, and unlock events, but did not track content type, screen brightness, or other qualitative aspects of usage. Daily screen time was defined as the total minutes of active device interaction within a calendar day, aggregated across all recorded events. Days without any recorded screen events were treated as missing and excluded. In this study, we analyzed screen time from the calendar day preceding each sleep night.

### Sleep assessment

We monitored sleep via the Ultrahuman Ring AIR, a finger-worn smart ring equipped with photoplethysmography (PPG), accelerometry, and skin-temperature sensors that enable continuous physiological monitoring during wakefulness and sleep. Sleep was assessed using the Ultrahuman sleep score, a composite metric derived from PPG-based heart rate variability, accelerometry-derived movement, and skin-temperature signals recorded during sleep [20,21]. The score ranges from 0 to 100, with higher values indicating better sleep quality; values below 21 were treated as sentinel values and excluded. Recovery score, a next-morning composite reflecting autonomic readiness derived from overnight HRV trends, resting heart rate, and respiratory rate, was extracted as a secondary outcome. Bedtime was detected algorithmically from the ring’s accelerometry and PPG signals and was used to define the pre-sleep window used in the timing analyses (i.e., the screen-time minutes within each of the 5 clock-hours preceding each user’s bedtime). Additional metrics analyzed included total sleep duration, deep sleep percentage, REM sleep percentage, and sleep efficiency.

### Statistical analysis

#### General approach

Continuous variables are reported as mean ± SEM unless otherwise noted. Categorical variables are reported as N (%) for user-level counts and n (%) for night-level counts. Because screen-time and sleep-score distributions were non-normal (Shapiro-Wilk p < 0.001), non-parametric methods were used for primary analyses. Effect sizes are reported as rank-biserial r for Mann-Whitney U tests and epsilon-squared for Kruskal-Wallis tests.

For per-metric Q1 vs Q5 comparisons (**Table 1A** user-level and 1B night-level), we used a normality-aware test scaffold: each metric was checked for distributional normality on each quintile arm with Shapiro-Wilk (W, two-sided p < 0.05 considered non-normal); if either arm was non-normal we used Mann-Whitney U with rank-biserial r as the effect size, otherwise we used Welch’s t-test with Cohen’s d. The chosen test for each metric is reported in the “Test” column of **Table 1**, and the matched effect-size column reports r or d accordingly.

Where we report a family of parallel tests in a single figure (e.g. the six sleep-architecture metrics in **Figure 1B**), p-values are adjusted with the Benjamini–Hochberg false-discovery-rate procedure across that family; the adjusted value is denoted q (so q < 0.05 controls the expected proportion of false positives at 5% across the family of tests). The forest plot in **Figure 1B** and the per-metric q-values reported in **Table 1** use this convention. Where only a single test is reported (e.g. an individual GLMM coefficient or a single transition-pair Wilcoxon), we report the raw p.

For ordered-stratum trend tests across screen-time quintiles (**Figure 1A** inset), we used the Jonckheere–Terpstra trend test, a non-parametric test for monotone ordering across an ordered set of groups; we report the standardised z statistic and the two-sided p-value.

The Ultrahuman sleep score is reported on a 0-100 scale; in this cohort, the between-user standard deviation of mean sleep score is 7.6 points (per-night SD ≈ 14.2 points). A 5-point difference therefore corresponds to roughly 0.7 between-user SD, and the 8.5-point DP–RCU gap reported in **Figure 3B** corresponds to roughly 1.1 between-user SD. For multi-metric forest plots (**Figure 1B**, **T**able 1) where the six sleep-architecture metrics differ in units (points, hours, percentages), we report Cohen’s d alongside raw mean differences so cross-metric magnitudes are comparable on a unit-free scale; rank-biserial r is reported in place of d wherever the test of record was Mann-Whitney U. The sleep score is a proprietary composite metric and we do not claim a published clinical-significance threshold for sleep-score point differences; readers should anchor sleep-score deltas to the cohort variability above.

### Mixed-effects model

A generalized linear mixed model (GLMM) with a random intercept for each user predicted nightly sleep score from daily screen time, with interaction terms for age group, sex, and day type (weekday vs weekend). The model was fitted on 337,371 nights from 2,958 users (reduced from 3,086 due to 128 users missing age or sex data). The intraclass correlation coefficient (ICC) was 0.269, indicating that approximately 27% of variance in nightly sleep score was attributable to between-user differences.

For night j of user i with daily-screen-time exposure ST_ij (in hours), nightly sleep score was modelled as a linear mixed model with a per-user random intercept u_i ∼ N(0, σ_u²):

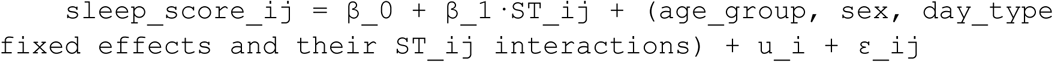

with age_group ∈ {<30, 30–39 (ref), 40–49, 50+}, sex ∈ {Male (ref), Female}, and day_type ∈ {Weekday (ref), Weekend}. Predictors were left in their natural units (screen time in hours, sleep score in points on the 0–100 scale) rather than z-scored, so the reported fixed-effect coefficients are directly interpretable as the expected change in sleep score per one-hour change in daily screen time and per one-category change in each moderator. The model was fit with statsmodels.mixedlm 0.14 in Python 3.12 using REML; the intra-class correlation ICC is reported in Results.

### Clustering and phenotype analysis

nTemporal usage phenotypes were identified by applying K-means clustering to L2-normalized 24-hour screen-use profiles. L2 normalization divides each user’s 24-hour screen-time vector by its Euclidean norm (the square root of the sum of squared hourly values), removing differences in total daily screen time and isolating the temporal shape of usage. Cluster-number selection was informed by per-user silhouette coefficients (**Supplementary Figure S2A**) and separation in sleep score across clusters (**Supplementary Figure S2C**). The K = 2 solution achieved the lowest absolute proportion of users with negative silhouette coefficients (6.2%); the K = 3 solution was a close second (7.1%). K = 3 was nonetheless adopted as the working solution because at K = 3 a small but distinct third cluster (the Round-the-Clock Users phenotype, characterized in the Results) emerged that did not exist at K = 2 and was the analytical reason for the cluster split. At K = 4 and above, additional clusters became progressively less distinguishable: at K = 4, two of the four clusters had indistinguishable sleep outcomes (Mann-Whitney U p = 1.0, r = 0.01). Longitudinal stability was assessed in users contributing at least four quarterly windows of ≥30 valid days each over a span of ≥12 months (N = 1,098), by clustering each user’s quarterly profile independently and computing a 3×3 transition matrix on the 878 users with a stable cluster across the final two quarters.

We acknowledge that mean silhouette at K = 3 (0.126) is below that at K = 2 (0.177); K = 3 was nonetheless adopted because K = 2 averaged a small but distinct round-the-clock phenotype into the broader population, removing a sleep-distinguishable cluster and reducing inter-cluster sleep separation. At K = 3, 7.1% of users had negative silhouette coefficients, predominantly within the RCU cluster. Restricting attention to those negative-silhouette RCU users, 94.6% lay closer to the LNU centroid than to the DP centroid, consistent with a continuous LNU → RCU gradient in nighttime screen share rather than ambiguity between RCU and the lowest-night-use phenotype.

Clustering was performed with scikit-learn 1.4 KMeans (n_init = 20, random_state = 42) on the L1-normalized 24-hour profiles. Silhouette coefficients were computed with scikit-learn (silhouette_samples), and the per-quarter longitudinal phenotype assignments used in the transition matrix (**Figure 3D**, **T**able 2) were generated by clustering each user-quarter profile independently and aligning each quarter’s three local centroids to the cohort-level reference centroids by maximum cosine similarity, with each reference cluster assigned to at most one local cluster per quarter. The reference centroids and the per-quarter centroids reproduced the same three behavioral shapes (a daytime-peaked profile, a late-night-peaked profile, and a flat round-the-clock profile), so the transition matrix is interpreted on the same phenotype scaffold across all quarters. A transition is defined as a change between the user’s cluster in the first quarter and the cluster they held in both of the last two recording quarters (a 1:2 baseline-to-follow-up split). Requiring the last-two-quarter cluster to be the same drops users whose cluster flipped in only a single quarter, which are most likely clustering noise rather than a sustained behavioural change.

### Blood biomarker analysis

The biomarker subcohort comprised 65 users from the matched cohort who had a phenotype assignment and at least one measured blood biomarker analyte on a contemporaneous Ultrahuman Blood Vision panel (DP n = 14, LNU n = 36, RCU n = 15). All Blood Vision panels included in this analysis were drawn from users who consented to research use of their de-identified biomarker data through the Ultrahuman onboarding process. For each of 28 established clinical cutoffs across glycaemic, lipid, hepatic, inflammatory, hormonal, and micronutrient domains we computed odds ratios comparing Round-the-Clock Users to Daytime Peakers using Fisher exact tests; a Haldane correction (+0.5 to all cells) was applied when any cell contained zero events, and 95% confidence intervals were estimated using Woolf’s method [25]. p-values were adjusted for multiple comparisons using the Benjamini–Hochberg procedure. Across the 65 users, panel coverage varied (median 7 analytes per user, range 1–28), so the per-cutoff analytical N varies between 11 (rarer markers such as IL-6 and prolactin) and 62 (common markers including HbA1c, the lipid panel, and total cholesterol). Per-cutoff and per-cluster denominators are displayed alongside each row of **Supplementary Figure S3** so every odds ratio can be read against its own denominator. Demographics for this subcohort are in **Table S2**.

### Software

Analyses were performed in Python 3.12 using pandas 2.2, scipy 1.12, statsmodels 0.14, and scikit-learn 1.4.

### Use of artificial intelligence

Artificial intelligence tools were used to assist with analysis scripting and editorial refinement. All code, statistical outputs, and interpretations were reviewed and verified by the authors, who take full responsibility for the accuracy and integrity of the work.

## Data Availability

Summary statistics and analysis outputs supporting the findings of this study are provided in the Supplementary Information. Individual-level data are not publicly available because they are proprietary to Ultrahuman Healthcare Pvt. Ltd. and subject to user privacy constraints. The proprietary algorithms underlying the Ultrahuman sleep score and recovery score are also protected as trade secrets and cannot be disclosed; the validation work supporting their use as objective sleep-quality measures is reported in the cited Ultrahuman validation studies [20, 21].

## Notes

### Competing Interest Statement

All authors are employees of Ultrahuman Healthcare Pvt Ltd, which develops and commercializes the Ultrahuman Ring described in this study. The authors declare no other competing interests.

### Funding Statement

The study was supported by Ultrahuman Healthcare Pvt. Ltd internal research funds.

### Author Declarations

This was a real-world, retrospective, observational study based on data derived from Ultrahuman platform users, conducted in accordance with Ultrahuman’s Terms of Use (https://ultrahumanapp.notion.site/TERMS-OF-USE-7e3c52cf6b4a47078a5989e90a82e326) and Privacy Policy (https://ultrahumanapp.notion.site/Ultrahuman-Privacy-Policy-12d72afc73e880519f1ce5fecfe58d7c), which permit the analysis of de-identified, aggregated data for scientific research. Participants consented through the onboarding process on the Ultrahuman platform and continued product use. No dietary, sleep, exercise, or other interventions were administered as part of this study: wearable-derived physiological signals and screen-time records were captured passively during routine product use, and blood biomarker results were obtained from contemporaneous Ultrahuman Blood Vision panels. All data were de-identified prior to analysis, and a separate team of analysts extracted the data, executed the computational pipelines, and reviewed the results to maintain analyst blinding.

