## Supplementary Information for "Pre-bed and nighttime screen use, beyond daily total, is inversely associated with sleep quality: a longitudinal study of 350,600 nights"

*for*

**Supplementary Figure & Table Legends**

**Supp Fig S1. GLMM predicted margins for sleep score across daily screen time, stratified by moderator.** A linear mixed model with random intercepts for users adjusted sleep score from daily screen time, with interaction terms for **(A)** age group, **(B)** sex, and **(C)** day type. Shaded bands represent 95% confidence intervals for the predicted margins. **(D)** Cohort-wide model fit (intercept, screen-time slope, ICC, sample size). **(E)** Per-stratum baseline and slope shifts relative to each reference category, with 95% confidence intervals. Model fitted on 337,371 nights; N = 2,958 users (reduced from the base cohort of 3,086 users due to 128 users missing age or sex data); ICC = 0.269.

**Supp Fig S2. K-means cluster-validation diagnostics on the 3,086-user matched cohort.** **(A)** Per-user silhouette plots for k = 2, 3, 4. **(B)** Percentage of users with negative silhouette (bars) and mean silhouette (line) across k ∈ {2…8}. **(C)** Mean sleep score by cluster for k = 2, 3, 4 with pairwise Mann-Whitney comparisons. **(D)** Per-k summary of negative-silhouette percentage, sleep-score spread (range across clusters), nighttime-share percentage of the highest-night-share cluster, and that cluster's mean sleep score. k = 3 was selected as the working choice based on the combination of silhouette structure, sleep-score separation across clusters, interpretability of the resulting phenotypes, and the requirement that all clusters be sleep-distinguishable.

**Supp Fig S3. Odds ratios for clinical biomarker abnormalities in Round-the-Clock Users vs Daytime Peakers.** Forest plot of odds ratios (OR) with 95% confidence intervals (Woolf method) for 28 established clinical cutoffs spanning glycaemic, lipid, hepatic, inflammatory, hormonal, and micronutrient domains, in an analytical subcohort of 65 users with both a phenotype assignment and at least one measured analyte on a contemporaneous Blood Vision panel (Daytime Peakers, n = 14; Late-Night Users, n = 36; Round-the-Clock Users, n = 15). A further 26 users had a panel on file but no individual analytes recorded against any of these 28 cutoffs and are excluded from this analysis (cohort-membership N = 91; analytical N = 65). Per-cutoff analytical N (shown in the DP / LNU / RCU columns alongside each row) varies between 11 and 62 because not every panel measured every analyte. OR > 1 indicates higher prevalence of abnormality in Round-the-Clock Users; OR < 1 indicates higher prevalence in Daytime Peakers. Haldane correction (+0.5 to all cells) was applied when any cell contained zero events and is flagged with a dagger (†) next to the affected odds ratios. Fisher’s exact test was used throughout; no individual comparison reached significance after FDR correction across the 28 cutoffs.

**Supp Table S1.** Participant-flow disposition table mirroring the cohort derivation in Methods. Columns report user count, paired screen-sleep night count where applicable, and the figure(s) each subcohort supports.

**Supp Table S2.** Per-phenotype demographic summary supporting **Figure 3C**: N, age (mean ± SD), sex split, BMI, peak/trough hour of screen-time profile, mean daily screen time, mean sleep score, and mean sleep duration for Daytime Peakers (DP), Late-Night Users (LNU), and Round-the-Clock Users (RCU).

**Supplementary Figures and Tables**

**Supp Fig S1**

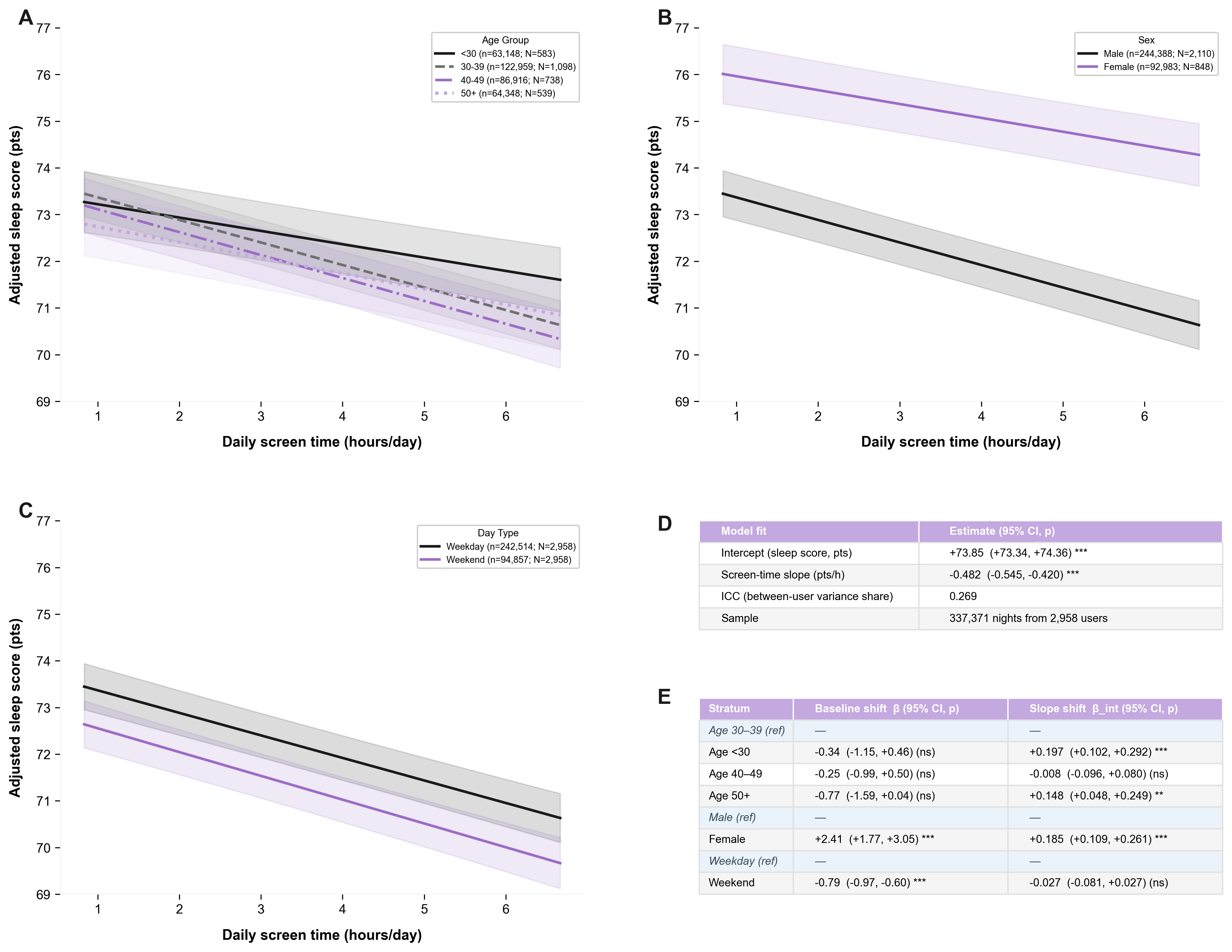

**Supp Fig S2**

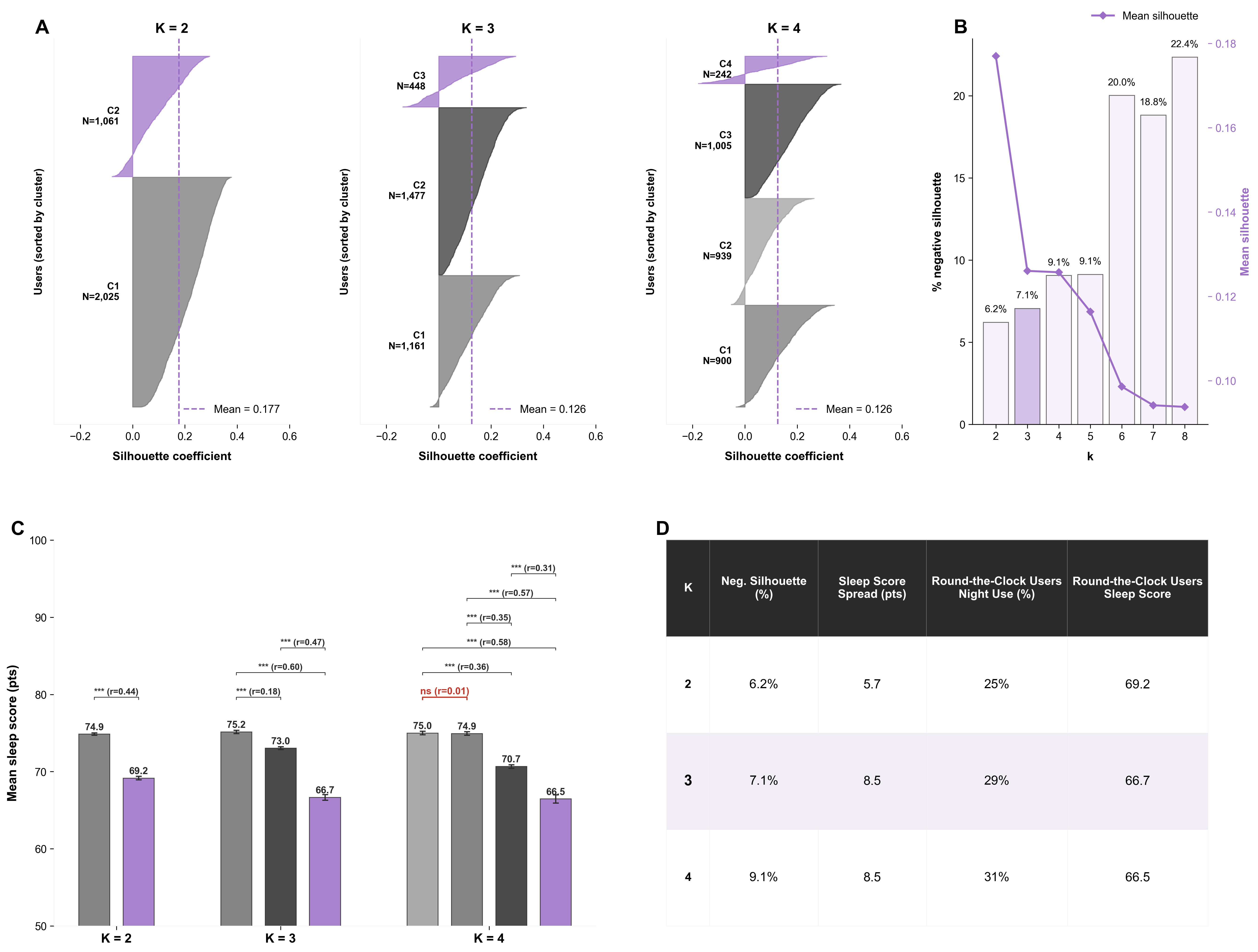

**Supp Fig S3**

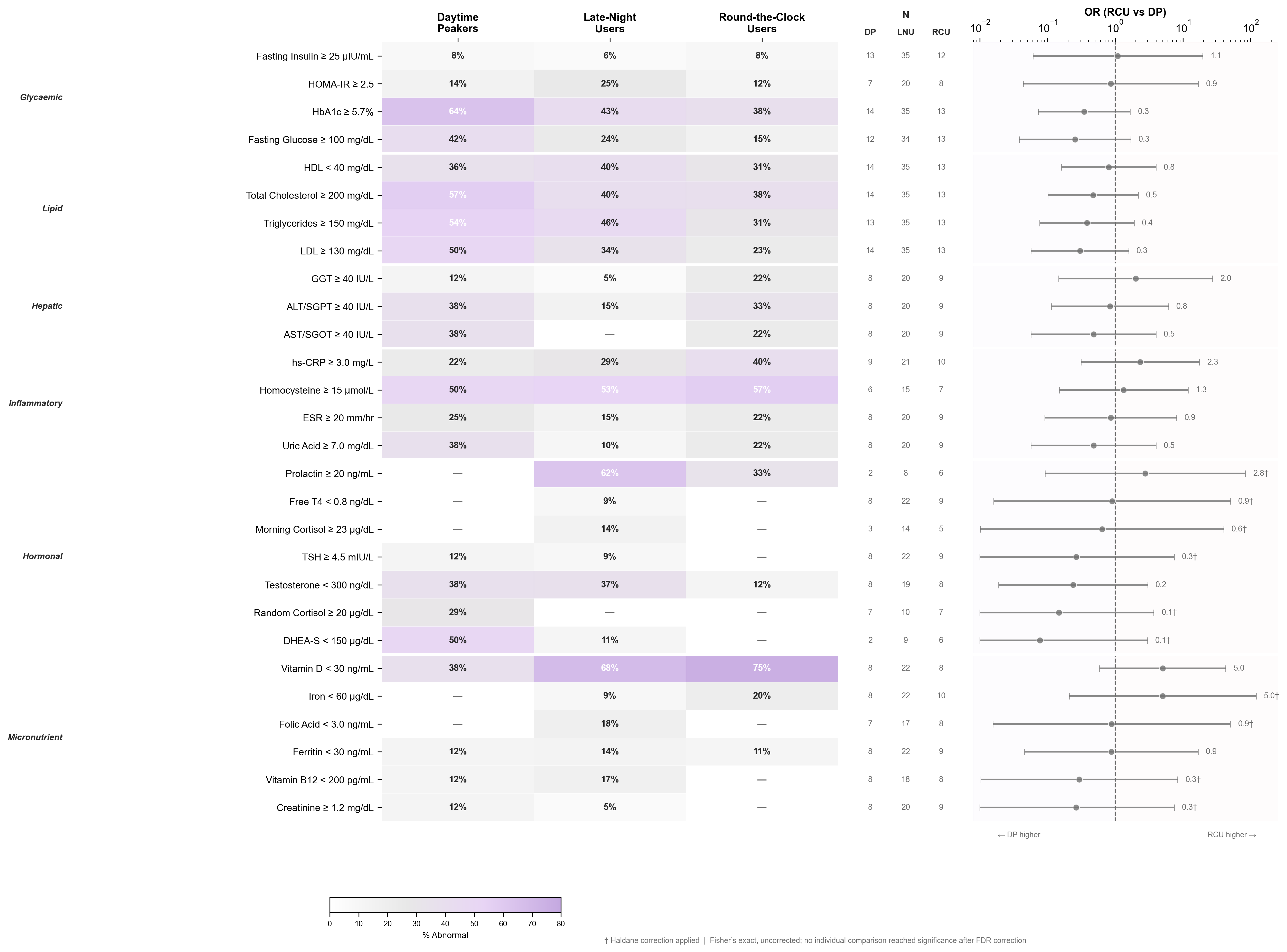

**Supp Table S1**

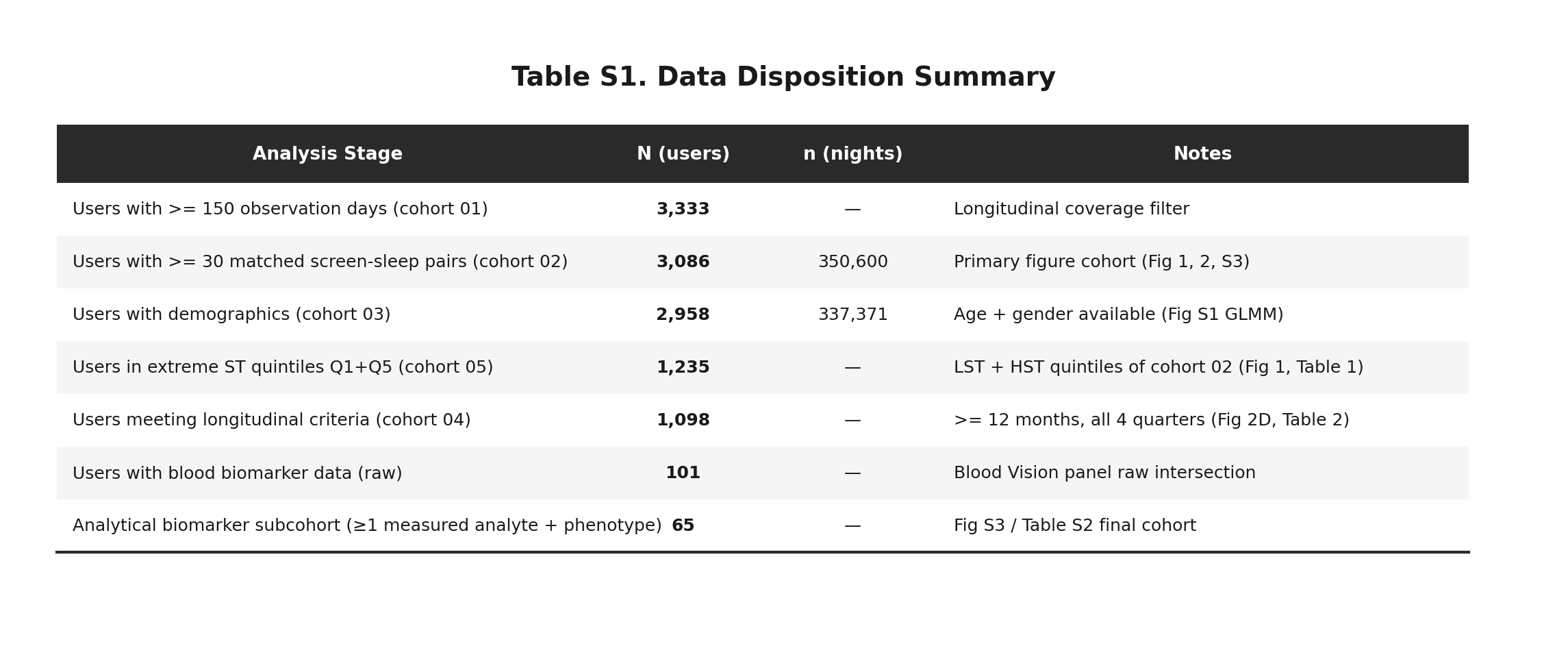

**Supp Table S2**

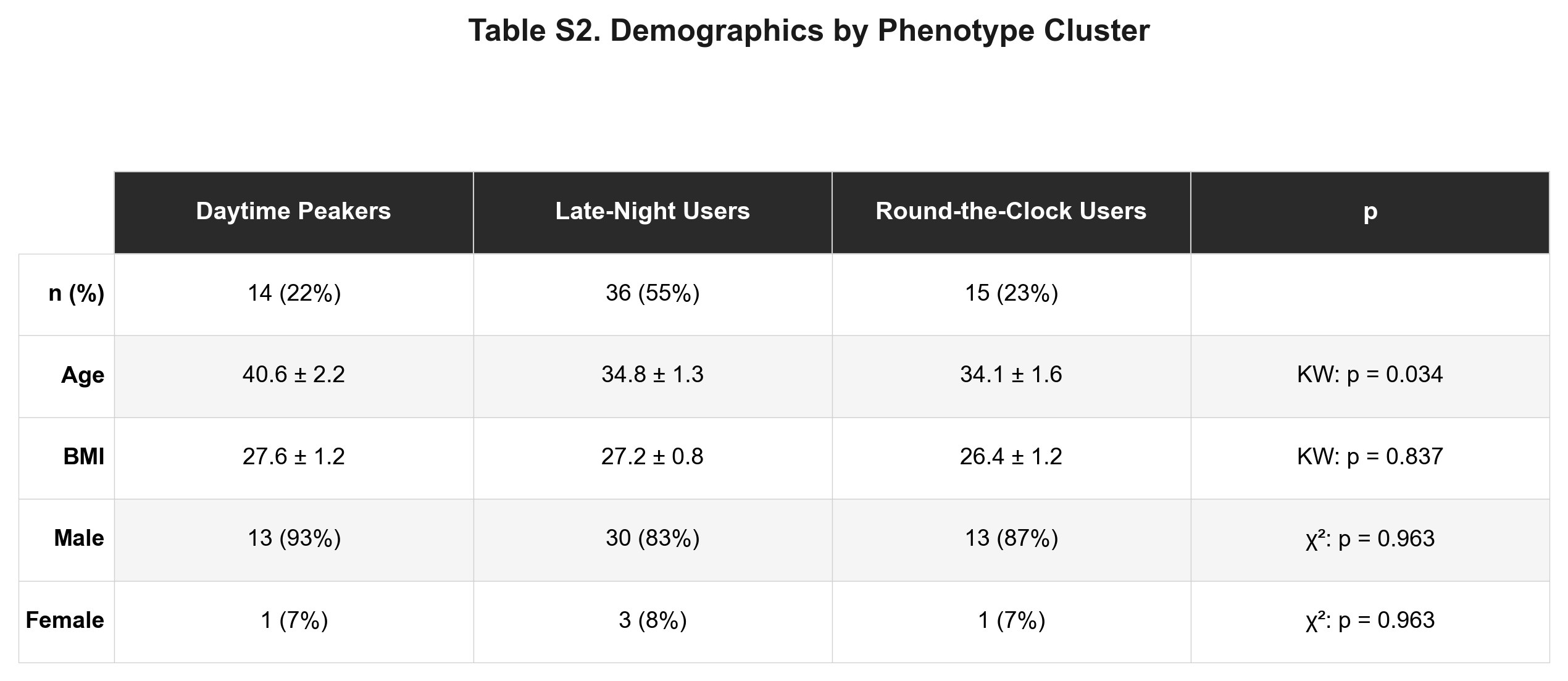
